# Psychological distress among people with probable COVID-19 infection: analysis of the UK Household Longitudinal Study

**DOI:** 10.1101/2020.11.24.20237909

**Authors:** Claire L Niedzwiedz, Michaela Benzeval, Kirsten Hainey, Alastair H Leyland, Srinivasa Vittal Katikireddi

## Abstract

Studies exploring the longer-term effects of experiencing COVID-19 infection on mental health are lacking. We explored the relationship between reporting probable COVID-19 symptoms in April 2020 and psychological distress (measured using the General Health Questionnaire) one, two, three, five and seven months later. Data were taken from the UK Household Longitudinal Study, a nationally representative household panel survey of UK adults. Elevated levels of psychological distress were found up to seven months after probable COVID-19, compared to participants with no likely infection. Associations were stronger among younger age groups and men. Further research into the psychological sequalae of COVID-19 is urgently needed.

## Introduction

Considerable concerns exist about persistent and debilitating symptoms experienced by people who have had COVID-19 infection, often referred to as ‘long-COVID’.(1) However, population-based data remain rare, with the majority of COVID-19 research focused on severe adverse physical consequences of acute disease. There is growing evidence that a substantial number of people experience persisting symptoms such as fatigue and chest pain months after infection.(2) While the mental health consequences of societal changes during the pandemic (including lockdown) have been extensively studied (3, 4), research on the impact of COVID-19 infection on mental health and the role of psychiatric symptoms in long-COVID is limited.

Research from previous coronavirus outbreaks demonstrates potential for psychiatric consequences of infection.(5) An initial study using administrative data from the US demonstrated that COVID-19 infection was associated with an increased incidence of psychiatric diagnoses (particularly anxiety and insomnia) in the following 14 to 90 days.(6) Studies have also demonstrated that COVID-19 survivors who received Emergency Department evaluation in Milan had high levels of depression and anxiety one month following discharge (7) and COVID-19 patients in Chongqing, China had more symptoms of stress, anxiety and depression compared to healthy and psychiatric patient controls (8). However, most existing studies have been limited by small, non-representative samples and those based on electronic health records may not capture individuals with more minor acute COVID-19 symptoms or those with less severe psychological problems that do not present to health services. Changes in healthcare seeking and treatment may lead to substantial potential bias in the context of a pandemic, where disruptions to health systems have been widespread. Data from surveys may therefore be better placed to capture impacts on mental health and need for intervention. To explore the relationship between COVID-19 infection and mental health in the UK context, we assessed associations between experiencing COVID-19 symptoms and changes in psychological distress in a representative longitudinal survey of adults.

## Methods

The UK Household Longitudinal Study (also referred to as Understanding Society) is a nationally representative longitudinal household panel survey, based on a clustered stratified probability sample of UK households, described in detail previously.(9) All adults (aged 16+ years) in chosen households are invited to participate. Data collection for each wave usually takes place over 24 months, with participants re-interviewed every year by online, face-to-face or telephone survey. We used pre-pandemic data from wave 9 (2017-19), which achieved a household response rate of over 80%.(9, 10) In response to the COVID-19 pandemic, additional waves of data were collected in 2020 via online survey during April (24^th^ to 30^th^ April), May (27^th^ May to 2^nd^ June), June (25^th^ June to 1^st^ July), July (24^th^ to 31^st^ July), September (24^th^ September to 1^st^ October) and November (24^th^ November to 1^st^ December).(11, 12) The response rate for the first COVID wave was 48.6% of those who took part at wave 9. (11, 13)

Psychological distress was measured at each wave (pre-pandemic in 2017-19 and during the pandemic at each wave as above) via the General Health Questionnaire 12 item instrument (GHQ-12). GHQ-12 assesses psychological distress and respondents reporting a score of four or more are likely experiencing symptoms to a clinically significant level.(3) Self-reported symptoms of cough, fever and anosmia allowed identification of individuals with probable COVID-19 infection in April 2020 (Supplementary Material). Of 41 people reporting hospitalisation for COVID-19 in April 2020, 34 were classified as probable COVID-19 according to our definition. We assessed associations between probable COVID-19 infection and psychological distress at one, two, three, five and seven months later using logistic regression, with models adjusted to account for pre-pandemic psychological distress (GHQ score) and other covariates (gender, age, ethnicity and longstanding illness or disability). We also conducted subgroup analyses by age group (under 45, 45-64, 65+ years) and gender (men, women). Analyses used inverse probability weights to adjust for non-response and standard errors were adjusted for the complex survey design. Missing data were excluded from analyses, but if participants were missing pre-pandemic wave 9 data responses from wave 10 were used if available. Statistical analyses were performed in Stata/MP 15.1.

The University of Essex Ethics Committee approved all data collection for the Understanding Society main survey and COVID waves, which were performed in accordance with the Declaration of Helsinki. All survey participants provided fully informed consent. No additional ethical approval was necessary for this secondary data analysis.

## Results

8.9% of participants experienced probable COVID-19 symptoms in April 2020 from a total of 12,492 participants. Psychological distress was more prevalent in May (27.4%, 95% CI: 25.9-28.9), reduced to 20.8% (95% CI: 19.4-22.3) in July, before increasing again to 26.5% (95% CI: 24.8-28.2) in November, corresponding with the UK winter lockdown. The characteristics of participants are described in Supplementary Table S1.

In comparison to participants without probable COVID-19 infection, psychological distress was more common at one (OR 1.39, 95% CI: 1.10-1.76), two (OR 1.38, 95% CI: 1.05-1.81), three (OR 1.31, 95% CI: 0.99-1.72), five (OR 1.42, 95% CI: 1.05-1.92) and seven (OR 1.47, 95% CI: 1.04-2.07) months after reporting COVID-19 symptoms in adjusted analyses (Table 1, Supplementary Table S2-S6). Subgroup analyses demonstrated stronger associations in men (OR=1.52 in May, 95% CI: 1.05-2.20) and younger adults (OR=1.51 in May, 95% CI: 1.07-2.13).

**Table 1:** Associations between probable COVID-19 infection in April 2020 and psychological distress (GHQ case) up to 7 months later stratified by age group and gender.

|  | 1 month later (May 2020) | 2 months later (June 2020) | 3 months later (July 2020) | 5 months later (September 2020) | 7 months later (November 2020) |
| --- | --- | --- | --- | --- | --- |
|  | OR<br>[95% CI] | OR<br>[95% CI] | OR<br>[95% CI] | OR<br>[95% CI] | OR<br>[95% CI] |
| **Overall** |  |  |  |  |  |
| No likely infection (reference) | 1 | 1 | 1 | 1 | 1 |
| Probable infection<sup>1</sup> | 1.39**<br>[1.10,1.76] | 1.38*<br>[1.05,1.81] | 1.31<br>[0.99,1.72] | 1.42*<br>[1.05,1.92] | 1.47*<br>[1.04,2.07] |
| N | 12,492 | 11,949 | 11,563 | 11,009 | 10,379 |
| **By gender** |  |  |  |  |  |
| Men | 1.52*<br>[1.05,2.20] | 1.57*<br>[1.04,2.37] | 1.43<br>[0.91,2.24] | 1.36<br>[0.88,2.12] | 1.77**<br>[1.20,2.61] |
| N | 5,138 | 4,908 | 4,759 | 4,556 | 4,283 |
| Women | 1.30<br>[0.98,1.75] | 1.31<br>[0.94,1.81] | 1.25<br>[0.86,1.82] | 1.46<br>[0.98,2.19] | 1.30<br>[0.82,2.07] |
| N | 7,279 | 6,969 | 6,730 | 6,379 | 6,022 |
| **By age group** |  |  |  |  |  |
| Under 45 years | 1.51*<br>[1.07,2.13] | 1.43<br>[0.91,2.26] | 1.20<br>[0.80,1.78] | 1.39<br>[0.89,2.18] | 1.74*<br>[1.02,2.95] |
| N | 3,426 | 3,169 | 2,996 | 2,735 | 2,481 |
| 45-64 years | 1.24<br>[0.81,1.90] | 1.39<br>[0.87,2.22] | 1.38<br>[0.85,2.25] | 1.53<br>[0.94,2.49] | 1.27<br>[0.67,2.38] |
| N | 5,279 | 5,045 | 4,924 | 4,724 | 4,485 |
| 65+ years | 1.16<br>[0.52,2.58] | 1.14<br>[0.63,2.04] | 1.33<br>[0.68,2.60] | 1.23<br>[0.52,2.92] | 1.14<br>[0.59,2.20] |
| N | 3,547 | 3,511 | 3,432 | 3,329 | 3,203 |

<sup>1</sup> Adjusted for age, sex, ethnicity, limiting long-standing illness, GHQ-12 at wave 9 (2017-19). CI=confidence interval; GHQ=General Health Questionnaire; OR=odds ratio.
\* p < 0.05, \*\* p < 0.01, \*\*\* p < 0.001
Results for each time point include participants who participated in Wave 9, COVID Wave A (April 2020) and the respective follow-up COVID Wave (May to November 2020)

## Discussion

Our study suggests that COVID-19 infection could lead to an increase in clinically significant psychological distress which persists months after infection and is additional to the mental health impacts of societal changes during the pandemic. Younger people and men with probable COVID-19 infection were more likely to report clinical levels of psychological distress compared to older age groups and women. Our findings add to the growing evidence that COVID-19 infection may have a direct impact on mental health (8, 14) that persists months after initial symptoms (6, 7). Study strengths include the longitudinal data over multiple points of follow-up during the pandemic, representative UK sample and accounting for pre-pandemic mental health to limit reverse causation. Important limitations include the ascertainment of COVID-19 infection, based on self-report only (not confirmed by a laboratory test) and the classification of probable COVID-19 infection at only one time point. However, misclassification may be more likely to result in under-estimation of any underlying association. Further research based on confirmed infection is required, as is research which takes into account any change in the symptoms of long-COVID. Triangulation of survey and administrative data (e.g. primary care and psychotropic prescribing records) would also be helpful to disentangle the discrepancy between likely clinical need and service use.

The potential adverse impacts of COVID-19 infection on mental health reinforces the benefits of minimising COVID-19 infection among the general population, not only in those at greatest risk of mortality. When considered alongside the mental health impacts generated by mitigation measures, there is potential for a high demand for mental health services resulting from the pandemic. Further research to examine the longer-term psychological sequelae of COVID-19 infection is urgently required.

## Data Availability

Understanding Society deidentified survey participant data are available through the UK Data Service (http://doi.org/10.5255/UKDA-SN-6614-14; http://doi.org/10.5255/UKDA-SN-8644-7). Researchers who would like to use Understanding Society need to register with the UK Data Service (https://ukdataservice.ac.uk/) before being allowed to download datasets.

## Author contributions

SVK and CLN conceived the idea for the study. CLN and SVK led the study design, with input from AHL. CLN and SVK conducted the analysis, with assistance from KH. SVK and CLN drafted the paper, with all authors critically revising the manuscript.

## Acknowledgements

We would like to thank the participants of the Understanding Society study. The Understanding Society COVID-19 study is funded by the Economic and Social Research Council (ES/K005146/1) and the Health Foundation (2076161). Fieldwork for the survey is carried out by Ipsos MORI and Kantar. Understanding Society is an initiative funded by the Economic and Social Research Council and various Government Departments, with scientific leadership by the Institute for Social and Economic Research, University of Essex.

## References

1. Yelin D, Wirtheim E, Vetter P, Kalil AC, Bruchfeld J, Runold M, et al. Long-term consequences of COVID-19: research needs. The Lancet Infectious Diseases. 2020; 20(10): 1115–7.

2. Kingstone T, Taylor AK, Donnell CA, Atherton H, Blane DN, Chew-Graham CA. Finding the ‘right’ GP: a qualitative study of the experiences of people with long-COVID. BJGP Open. 2020: bjgpopen20X101143.

3. Niedzwiedz CL, Green MJ, Benzeval M, Campbell D, Craig P, Demou E, et al. Mental health and health behaviours before and during the initial phase of the COVID-19 lockdown: longitudinal analyses of the UK Household Longitudinal Study. Journal of Epidemiology and Community Health. 2020: jech-2020–215060.

4. O’Connor RC, Wetherall K, Cleare S, McClelland H, Melson AJ, Niedzwiedz CL, et al. Mental health and wellbeing during the COVID-19 pandemic: longitudinal analyses of adults in the UK COVID-19 Mental Health & Wellbeing study. The British Journal of Psychiatry. 2020: 1–17.

5. Rogers JP, Chesney E, Oliver D, Pollak TA, McGuire P, Fusar-Poli P, et al. Psychiatric and neuropsychiatric presentations associated with severe coronavirus infections: a systematic review and meta-analysis with comparison to the COVID-19 pandemic. The Lancet Psychiatry. 2020; 7(7): 611–27.

6. Taquet M, Luciano S, Geddes JR, Harrison PJ. Bidirectional associations between COVID-19 and psychiatric disorder: retrospective cohort studies of 62 354 COVID-19 cases in the USA. The Lancet Psychiatry.

7. Mazza MG, De Lorenzo R, Conte C, Poletti S, Vai B, Bollettini I, et al. Anxiety and depression in COVID-19 survivors: Role of inflammatory and clinical predictors. Brain, Behavior, and Immunity. 2020; 89: 594–600.

8. Hao F, Tam W, Hu X, Tan W, Jiang L, Jiang X, et al. A quantitative and qualitative study on the neuropsychiatric sequelae of acutely ill COVID-19 inpatients in isolation facilities. Translational Psychiatry. 2020; 10(1): 355.

9. Mulick A, Walker J, Puntis S, Burke K, Symeonides S, Gourley C, et al. Does depression treatment improve the survival of depressed patients with cancer? A long-term follow-up of participants in the SMaRT Oncology-2 and 3 trials. The Lancet Psychiatry.

10. Kanani R, Davies EA, Hanchett N, Jack RH. The association of mood disorders with breast cancer survival: an investigation of linked cancer registration and hospital admission data for South East England. Psycho-Oncology. 2016; 25(1): 19–27.

11. Wang SM, Chang JC, Weng SC, Yeh MK, Lee CS. Risk of suicide within 1 year of cancer diagnosis. International journal of cancer. 2018; 142(10): 1986–93.

12. Osazuwa-Peters N, Simpson MC, Zhao L, Boakye EA, Olomukoro SI, Deshields T, et al. Suicide risk among cancer survivors: Head and neck versus other cancers. Cancer. 2018; 124(20): 4072–9.

13. Benzeval M, Burton J, Crossley TF, Fisher P, Jäckle A, Low H, et al. The Idiosyncratic Impact of an Aggregate Shock: The Distributional Consequences of COVID-19. Available at SSRN 3615691. 2020.

14. Wang C, Pan R, Wan X, Tan Y, Xu L, McIntyre RS, et al. A longitudinal study on the mental health of general population during the COVID-19 epidemic in China. Brain Behav Immun. 2020; 87: 40–8.

