## Supplementary Material for "Psychological distress among people with probable COVID-19 infection: analysis of the UK Household Longitudinal Study"

Classification of probable COVID-19 infection

The following questions were asked:

Have you experienced symptoms that could be caused by coronavirus (COVID-19)? (Yes/No).

If the respondent answered ‘yes’ they were then asked the following:

Which of the following symptoms have you had?

Please select all that apply.

1. High temperature

2. A new continuous cough

3. Shortness of breath or trouble breathing

4. Runny or stuffy nose

5. Muscle or body aches

6. Headaches

7. Sore throat

8. Fatigue

9. Diarrhoea/Digestive issues/Upset stomach

10. Loss of sense of smell or taste

11. None of these

Based on the symptoms included in the COVID-19 case definition from [NHS England](https://www.nhs.uk/conditions/coronavirus-covid-19/testing-and-tracing/get-a-test-to-check-if-you-have-coronavirus/), we classified all respondents who had any one of responses 1, 2 or 10 as probable infection.

**Supplementary Table S1: Characteristics of participants included in the sample in April 2020 and at one, two, three, five and seven month intervals (weighted)**

|  | **1 month later (April and May 2020)** | **2 months later (April and June 2020)** | **3 months later (April and July 2020)** | **5 months later (April and September 2020)** | **7 months later (April and November 2020)** |
| --- | --- | --- | --- | --- | --- |
|  | % | % | % | % | % |
| ***Gender*** |  |  |  |  |  |
| Men | 47.2 | 47.1 | 47.6 | 46.9 | 46.7 |
| Women | 52.8 | 52.9 | 52.4 | 53.1 | 53.3 |
| ***Age group*** |  |  |  |  |  |
| Under 25 years | 9.6 | 9.9 | 9.2 | 9.9 | 9.5 |
| 25-44 years | 28.6 | 28.1 | 28.4 | 28.5 | 27.9 |
| 45-64 years | 37.9 | 37.8 | 37.9 | 37.8 | 38.4 |
| 65+ years | 23.9 | 24.2 | 24.5 | 23.9 | 24.1 |
| ***Ethnicity*** |  |  |  |  |  |
| White | 92.5 | 92.3 | 93.0 | 91.8 | 92.3 |
| Asian | 4.0 | 4.0 | 3.8 | 4.2 | 3.9 |
| Black | 1.4 | 1.6 | 1.3 | 1.8 | 1.7 |
| Mixed | 1.6 | 1.7 | 1.4 | 1.7 | 1.5 |
| Other | 0.5 | 0.5 | 0.5 | 0.6 | 0.6 |
| ***Longstanding illness or disability*** | 35.7 | 35.0 | 36.3 | 36.2 | 35.9 |
| ***Probable COVID-19 in April 2020*** | 8.9 | 8.6 | 8.8 | 8.8 | 8.8 |
| ***Psychological distress,***  ***GHQ case at follow-up***  (%, 95% CI) | 27.4 (25.9-28.9) | 26.0 (24.5-27.6) | 20.8 (19.4-22.3) | 21.0 (19.6-22.4) | 26.5 (24.8-28.2) |
| ***Psychological distress, GHQ score at Wave 9 (2017-19)*** (mean, 95% CI) | 1.85 (1.73-1.97) | 1.84 (1.72-1.97) | 1.82 (1.70-1.94) | 1.85 (1.72-1.97) | 1.86 (1.72-2.00) |
| N | 12,492 | 11,949 | 11,563 | 11,009 | 10,379 |

CI=confidence interval; GHQ=General Health Questionnaire

**Supplementary Table S2: Full results of logistic regression models one month after COVID-19 symptom reporting**

|  | **All** | **Men** | **Women** | **Under 45 years** | **45-64 years** | **65+ years** |
| --- | --- | --- | --- | --- | --- | --- |
|  | **OR**  **[95% CI]** | **OR**  **[95% CI]** | **OR**  **[95% CI]** | **OR**  **[95% CI]** | **OR**  **[95% CI]** | **OR**  **[95% CI]** |
| **Outcome: GHQ case (May 2020)** |  |  |  |  |  |  |
| **Probable COVID-19 infection:** No (reference) |  |  |  |  |  |  |
| Yes | 1.39^**^ [1.10,1.76] | 1.52^*^ [1.05,2.20] | 1.30 [0.98,1.75] | 1.51^*^ [1.07,2.13] | 1.24 [0.81,1.90] | 1.16 [0.52,2.58] |
| **GHQ score at Wave 9 (2017-19) (continuous)** | 1.23^***^ [1.20,1.27] | 1.29^***^ [1.24,1.35] | 1.20^***^ [1.16,1.23] | 1.20^***^ [1.15,1.25] | 1.24^***^ [1.20,1.28] | 1.30^***^ [1.19,1.41] |
| **Age group:** Under 25 years | 2.95^***^ [2.09,4.17] | 2.95^***^ [1.67,5.19] | 2.89^***^ [1.96,4.26] |  |  |  |
| 25 to 44 | 2.28^***^ [1.78,2.92] | 2.65^***^ [1.82,3.85] | 2.02^***^ [1.52,2.68] |  |  |  |
| 45 to 64 | 1.38^**^ [1.11,1.71] | 1.30 [0.99,1.70] | 1.41^**^ [1.10,1.80] |  |  |  |
| 65+ (reference) |  |  |  |  |  |  |
| **Sex:** Male (reference) |  |  |  |  |  |  |
| Female | 1.52^***^ [1.29,1.79] |  |  | 1.37^*^ [1.01,1.86] | 1.67^***^ [1.34,2.08] | 1.65^*^ [1.05,2.59] |
| **Ethnicity:** White (reference) |  |  |  |  |  |  |
| Asian | 0.83 [0.56,1.23] | 1.19 [0.74,1.91] | 0.60^*^ [0.37,0.95] | 0.75 [0.38,1.49] | 0.93 [0.61,1.41] | 1.13 [0.38,3.34] |
| Black | 0.43 [0.11,1.59] | 1.07 [0.47,2.47] | 0.18^*^ [0.04,0.86] | 0.49 [0.13,1.83] | 0.40 [0.09,1.78] | 0.12^*^ [0.02,0.64] |
| Mixed | 1.45 [0.61,3.41] | 1.33 [0.35,5.08] | 1.65 [0.54,5.08] | 1.43 [0.51,3.97] | 0.98 [0.33,2.95] | 8.51 [0.45,160.96] |
| Other | 0.84 [0.35,2.03] | 0.60 [0.17,2.17] | 1.16 [0.31,4.37] | 0.79 [0.34,1.84] | 1.19 [0.33,4.34] | 0.83 [0.46,1.49] |
| **Long-standing illness or disability:** No (reference) |  |  |  |  |  |  |
| Yes | 1.38^***^ [1.16,1.64] | 1.63^***^ [1.25,2.12] | 1.24^*^ [1.01,1.52] | 1.18 [0.82,1.71] | 1.69^***^ [1.33,2.13] | 1.37 [0.83,2.25] |
| **Age (continuous)** |  |  |  | 0.98^*^ [0.97,1.00] | 0.99 [0.97,1.01] | 0.98 [0.94,1.02] |
| **N** | 12492 | 5138 | 7279 | 3426 | 5279 | 3547 |

CI=confidence interval; GHQ=General Health Questionnaire; OR=odds ratio. * *p* < 0.05, ** *p* < 0.01, *** *p* < 0.001

Analysis used inverse probability weights and standard errors adjusted for the complex survey design. Missing data excluded from analyses.

**Supplementary Table S3: Full results of logistic regression models two months after COVID-19 symptom reporting**

|  | **All** | **Men** | **Women** | **Under 45 years** | **45-64 years** | **65+ years** |
| --- | --- | --- | --- | --- | --- | --- |
|  | **OR**  **[95% CI]** | **OR**  **[95% CI]** | **OR**  **[95% CI]** | **OR**  **[95% CI]** | **OR**  **[95% CI]** | **OR**  **[95% CI]** |
| **Outcome: GHQ case (June 2020)** | **All** | **Men** | **Women** | **Under 45 years** | **45-64 years** | **Over 65 years** |
| **Probable COVID-19 infection:** No (reference) |  |  |  |  |  |  |
| Yes | 1.38^*^ [1.05,1.81] | 1.57^*^ [1.04,2.37] | 1.31 [0.94,1.81] | 1.43 [0.91,2.26] | 1.39 [0.87,2.22] | 1.14 [0.63,2.04] |
| **GHQ score at Wave 9 (2017-19) (continuous)** | 1.24^***^ [1.20,1.28] | 1.31^***^ [1.26,1.37] | 1.20^***^ [1.15,1.24] | 1.23^***^ [1.18,1.29] | 1.24^***^ [1.20,1.28] | 1.25^***^ [1.16,1.34] |
| **Age group:** Under 25 years | 2.15^***^ [1.50,3.07] | 1.87^*^ [1.09,3.23] | 2.27^***^ [1.48,3.47] |  |  |  |
| 25 to 44 | 1.93^***^ [1.48,2.51] | 2.56^***^ [1.78,3.69] | 1.57^**^ [1.18,2.10] |  |  |  |
| 45 to 64 | 1.26 [0.98,1.61] | 1.54^**^ [1.18,2.02] | 1.09 [0.84,1.41] |  |  |  |
| 65+ (reference) |  |  |  |  |  |  |
| **Sex:** Male (reference) |  |  |  |  |  |  |
| Female | 1.49^***^ [1.25,1.77] |  |  | 1.45^*^ [1.07,1.97] | 1.32^*^ [1.01,1.73] | 2.05^***^ [1.35,3.09] |
| **Ethnicity:** White (reference) |  |  |  |  |  |  |
| Asian | 1.13 [0.80,1.58] | 1.42 [0.88,2.30] | 0.99 [0.61,1.62] | 1.00 [0.58,1.72] | 1.21 [0.79,1.86] | 1.66 [0.97,2.83] |
| Black | 0.88 [0.26,2.99] | 1.38 [0.60,3.16] | 0.59 [0.10,3.48] | 0.95 [0.25,3.51] | 0.92 [0.23,3.67] | 0.13^**^ [0.03,0.58] |
| Mixed | 1.99 [0.85,4.66] | 2.03 [0.93,4.45] | 2.05 [0.75,5.62] | 2.13 [0.76,5.96] | 1.58 [0.55,4.50] | 0.43 [0.04,4.14] |
| Other | 0.47 [0.11,1.96] | 0.83 [0.25,2.71] | 0.45 [0.07,3.12] | 0.68 [0.23,2.04] | 0.18 [0.00,9.28] | 0.89 [0.54,1.46] |
| **Long-standing illness or disability:** No (reference) |  |  |  |  |  |  |
| Yes | 1.27^**^ [1.06,1.52] | 1.46^**^ [1.10,1.93] | 1.16 [0.94,1.42] | 1.08 [0.73,1.59] | 1.48^**^ [1.15,1.91] | 1.43 [0.92,2.24] |
| **Age (continuous)** |  |  |  | 0.99 [0.97,1.01] | 0.98^*^ [0.96,1.00] | 0.99 [0.95,1.02] |
| **N** | 11949 | 4908 | 6969 | 3169 | 5045 | 3511 |

CI=confidence interval; GHQ=General Health Questionnaire; OR=odds ratio. * *p* < 0.05, ** *p* < 0.01, *** *p* < 0.001

Analysis used inverse probability weights and standard errors adjusted for the complex survey design. Missing data excluded from analyses.

**Supplementary Table S4: Full results of logistic regression models three months after COVID-19 symptom reporting**

|  | **All** | **Men** | **Women** | **Under 45 years** | **45-64 years** | **65+ years** |
| --- | --- | --- | --- | --- | --- | --- |
|  | **OR**  **[95% CI]** | **OR**  **[95% CI]** | **OR**  **[95% CI]** | **OR**  **[95% CI]** | **OR**  **[95% CI]** | **OR**  **[95% CI]** |
| **Outcome: GHQ case (July 2020)** | **All** | **Men** | **Women** | **Under 45 years** | **45-64 years** | **Over 65 years** |
| **Probable COVID-19 infection:** No (reference) |  |  |  |  |  |  |
| Yes | 1.31 [0.99,1.72] | 1.43 [0.91,2.24] | 1.25 [0.86,1.82] | 1.20 [0.80,1.78] | 1.38 [0.85,2.25] | 1.33 [0.68,2.60] |
| **GHQ score at Wave 9 (2017-19) (continuous)** | 1.22^***^ [1.19,1.26] | 1.29^***^ [1.23,1.35] | 1.19^***^ [1.15,1.23] | 1.19^***^ [1.14,1.24] | 1.23^***^ [1.19,1.28] | 1.32^***^ [1.23,1.42] |
| **Age group:** Under 25 years | 1.81^**^ [1.21,2.71] | 1.76 [0.99,3.11] | 1.79^*^ [1.12,2.87] |  |  |  |
| 25 to 44 | 1.94^***^ [1.46,2.56] | 3.09^***^ [1.99,4.80] | 1.35 [0.99,1.83] |  |  |  |
| 45 to 64 | 1.41^**^ [1.10,1.80] | 1.48^*^ [1.09,2.02] | 1.33^*^ [1.00,1.76] |  |  |  |
| 65+ (reference) |  |  |  |  |  |  |
| **Sex:** Male (reference) |  |  |  |  |  |  |
| Female | 1.26^*^ [1.05,1.51] |  |  | 0.98 [0.68,1.41] | 1.52^***^ [1.19,1.94] | 1.69^*^ [1.01,2.81] |
| **Ethnicity:** White (reference) |  |  |  |  |  |  |
| Asian | 1.03 [0.66,1.61] | 0.88 [0.52,1.49] | 1.18 [0.64,2.18] | 1.01 [0.51,2.00] | 0.81 [0.52,1.27] | 1.19 [0.33,4.27] |
| Black | 0.76 [0.21,2.68] | 1.41 [0.61,3.26] | 0.61 [0.11,3.41] | 1.32 [0.38,4.61] | 0.45 [0.11,1.83] | 0.30^***^ [0.20,0.44] |
| Mixed | 1.82 [0.67,4.97] | 2.59^**^ [1.33,5.05] | 1.33 [0.62,2.83] | 1.83 [0.35,9.65] | 1.57 [0.56,4.36] | 0.91 [0.28,2.97] |
| Other | 0.54 [0.12,2.42] | 0.69 [0.16,3.06] | 0.54 [0.05,5.54] | 0.90 [0.30,2.70] | 0.08 [0.00,5.78] | 1.27 [0.68,2.36] |
| **Long-standing illness or disability:** No (reference) |  |  |  |  |  |  |
| Yes | 1.31^**^ [1.09,1.58] | 1.55^**^ [1.13,2.12] | 1.18 [0.95,1.46] | 1.07 [0.72,1.59] | 1.70^***^ [1.32,2.18] | 1.37 [0.80,2.36] |
| **Age (continuous)** |  |  |  | 1.00 [0.98,1.02] | 0.97^*^ [0.95,1.00] | 0.97 [0.93,1.02] |
| **N** | 11563 | 4759 | 6730 | 2996 | 4924 | 3432 |

CI=confidence interval; GHQ=General Health Questionnaire; OR=odds ratio. * *p* < 0.05, ** *p* < 0.01, *** *p* < 0.001

Analysis used inverse probability weights and standard errors adjusted for the complex survey design. Missing data excluded from analyses.

**Supplementary Table S5: Full results of logistic regression models five months after COVID-19 symptom reporting**

|  | **All** | **Men** | **Women** | **Under 45 years** | **45-64 years** | **65+ years** |
| --- | --- | --- | --- | --- | --- | --- |
|  | **OR**  **[95% CI]** | **OR**  **[95% CI]** | **OR**  **[95% CI]** | **OR**  **[95% CI]** | **OR**  **[95% CI]** | **OR**  **[95% CI]** |
| **Outcome: GHQ case (September 2020)** | **All** | **Men** | **Women** | **Under 45 years** | **45-64 years** | **Over 65 years** |
| **Probable COVID-19 infection:** No (reference) |  |  |  |  |  |  |
| Yes | 1.42^*^ [1.05,1.92] | 1.36 [0.88,2.12] | 1.46 [0.98,2.19] | 1.39 [0.89,2.18] | 1.53 [0.94,2.49] | 1.23 [0.52,2.92] |
| **GHQ score at Wave 9 (2017-19) (continuous)** | 1.25^***^ [1.21,1.28] | 1.30^***^ [1.24,1.36] | 1.21^***^ [1.17,1.26] | 1.24^***^ [1.19,1.30] | 1.23^***^ [1.18,1.27] | 1.31^**^ [1.11,1.54] |
| **Age group:** Under 25 years | 1.55^*^ [1.02,2.35] | 2.12^*^ [1.18,3.80] | 1.29 [0.75,2.20] |  |  |  |
| 25 to 44 | 1.55^**^ [1.17,2.04] | 1.86^**^ [1.29,2.70] | 1.35 [0.98,1.87] |  |  |  |
| 45 to 64 | 1.34^*^ [1.04,1.73] | 1.47^*^ [1.07,2.02] | 1.26 [0.95,1.69] |  |  |  |
| 65+ (reference) |  |  |  |  |  |  |
| **Sex:** Male (reference) |  |  |  |  |  |  |
| Female | 1.51^***^ [1.25,1.82] |  |  | 1.29 [0.93,1.80] | 1.59^***^ [1.24,2.05] | 1.94^*^ [1.16,3.22] |
| **Ethnicity:** White (reference) |  |  |  |  |  |  |
| Asian | 0.87 [0.60,1.27] | 0.77 [0.42,1.40] | 0.94 [0.58,1.53] | 0.80 [0.48,1.34] | 0.78 [0.50,1.20] | 2.01 [0.73,5.56] |
| Black | 0.42 [0.11,1.60] | 0.93 [0.39,2.26] | 0.20 [0.03,1.52] | 0.45 [0.06,3.41] | 0.44 [0.12,1.55] | 0.16^***^ [0.06,0.40] |
| Mixed | 0.58 [0.31,1.09] | 0.82 [0.28,2.41] | 0.46^*^ [0.23,0.92] | 0.37^***^ [0.21,0.65] | 1.31 [0.33,5.13] | 1.52 [0.32,7.12] |
| Other | 0.46 [0.08,2.61] | 0.99 [0.23,4.21] | 0.35 [0.03,4.42] | 0.94 [0.25,3.50] | 0.04 [0.00,2.28] | 1.03 [0.58,1.82] |
| **Long-standing illness or disability:** No (reference) |  |  |  |  |  |  |
| Yes | 1.24^*^ [1.03,1.50] | 1.39^*^ [1.02,1.89] | 1.18 [0.95,1.47] | 1.05 [0.74,1.49] | 1.45^**^ [1.13,1.87] | 1.32 [0.79,2.21] |
| **Age (continuous)** |  |  |  | 0.99 [0.97,1.02] | 0.98 [0.96,1.01] | 0.99 [0.95,1.04] |
| **N** | 11009 | 4556 | 6379 | 2735 | 4724 | 3329 |

CI=confidence interval; GHQ=General Health Questionnaire; OR=odds ratio. * *p* < 0.05, ** *p* < 0.01, *** *p* < 0.001

Analysis used inverse probability weights and standard errors adjusted for the complex survey design. Missing data excluded from analyses.

**Supplementary Table S6: Full results of logistic regression models seven months after COVID-19 symptom reporting**

|  | **All** | **Men** | **Women** | **Under 45 years** | **45-64 years** | **65+ years** |
| --- | --- | --- | --- | --- | --- | --- |
|  | **OR**  **[95% CI]** | **OR**  **[95% CI]** | **OR**  **[95% CI]** | **OR**  **[95% CI]** | **OR**  **[95% CI]** | **OR**  **[95% CI]** |
| **Outcome: GHQ case (November 2020)** | **All** | **Men** | **Women** | **Under 45 years** | **45-64 years** | **Over 65 years** |
| **Probable COVID-19 infection:** No (reference) |  |  |  |  |  |  |
| Yes | 1.47^*^ [1.04,2.07] | 1.77^**^ [1.20,2.61] | 1.30 [0.82,2.07] | 1.74^*^ [1.02,2.95] | 1.27 [0.67,2.38] | 1.14 [0.59,2.20] |
| **GHQ score at Wave 9 (2017-19) (continuous)** | 1.21^***^ [1.17,1.25] | 1.27^***^ [1.20,1.33] | 1.17^***^ [1.12,1.22] | 1.19^***^ [1.13,1.25] | 1.21^***^ [1.16,1.27] | 1.25^***^ [1.16,1.36] |
| **Age group:** Under 25 years | 2.01^***^ [1.37,2.96] | 1.89 [0.99,3.59] | 2.03^**^ [1.30,3.15] |  |  |  |
| 25 to 44 | 1.82^***^ [1.39,2.37] | 1.79^**^ [1.23,2.60] | 1.80^***^ [1.31,2.49] |  |  |  |
| 45 to 64 | 1.21 [0.95,1.54] | 1.21 [0.89,1.63] | 1.20 [0.91,1.60] |  |  |  |
| 65+ (reference) |  |  |  |  |  |  |
| **Sex:** Male (reference) |  |  |  |  |  |  |
| Female | 1.61^***^ [1.34,1.93] |  |  | 1.59^**^ [1.15,2.19] | 1.57^***^ [1.23,2.00] | 1.71^*^ [1.11,2.64] |
| **Ethnicity:** White (reference) |  |  |  |  |  |  |
| Asian | 0.68 [0.44,1.07] | 0.71 [0.42,1.21] | 0.64 [0.41,1.02] | 0.61 [0.34,1.10] | 0.62 [0.33,1.17] | 1.82 [0.59,5.64] |
| Black | 0.32^*^ [0.11,0.95] | 0.55^**^ [0.37,0.84] | 0.23^*^ [0.06,0.81] | 0.43^*^ [0.21,0.88] | 0.24^***^ [0.13,0.44] | 0.05^***^ [0.02,0.17] |
| Mixed | 0.96 [0.51,1.80] | 0.88 [0.39,2.01] | 1.02 [0.37,2.80] | 1.02 [0.64,1.64] | 0.66 [0.23,1.88] | 0.90 [0.13,6.29] |
| Other | 0.65 [0.18,2.37] | 0.79 [0.33,1.91] | 0.67 [0.11,4.20] | 1.26 [0.20,7.90] | 0.10 [0.00,3.96] | 0.65 [0.36,1.17] |
| **Long-standing illness or disability:** No (reference) |  |  |  |  |  |  |
| Yes | 1.37^**^ [1.13,1.66] | 1.39^*^ [1.05,1.84] | 1.35^*^ [1.07,1.70] | 1.13 [0.75,1.68] | 1.68^***^ [1.26,2.25] | 1.47 [0.91,2.38] |
| **Age (continuous)** |  |  |  | 0.99 [0.97,1.00] | 0.97^**^ [0.95,0.99] | 1.01 [0.96,1.05] |
| **N** | 10379 | 4283 | 6022 | 2481 | 4485 | 3203 |

CI=confidence interval; GHQ=General Health Questionnaire; OR=odds ratio. * *p* < 0.05, ** *p* < 0.01, *** *p* < 0.001

Analysis used inverse probability weights and standard errors adjusted for the complex survey design. Missing data excluded from analyses.
